# Intrinsic growth rules of patients infected, dead and recovered with 2019 novel coronavirus in mainland China

**DOI:** 10.1101/2020.02.23.20024802

**Authors:** Chuanliang Han, Yimeng Liu, Saini Yang

## Abstract

An outbreak of a novel coronavirus (SARS-CoV-2)-infected pneumonia (COVID-19) was first diagnosed in Wuhan, China, in December 2019 and then spread rapidly to other regions. We collected the time series data of the cumulative number of confirmed infected, dead, and cured cases from the health commissions in 31 provinces in mainland China. A descriptive model in a logistic form was formulated to infer the intrinsic epidemic rules of COVID-19, which illustrates robustness spatially and temporally. Our model is robust (*R*^*2*^>0.95) to depict the intrinsic growth rule for the cumulative number of confirmed infected, dead, and cured cases in 31 provinces in mainland China. Furthermore, we compared the intrinsic epidemic rules of COVID-19 in Hubei with that of severe acute respiratory syndrome (SARS) in Beijing, which was obtained from the Ministry of Public Health of China in 2003. We found that the infected case is the earliest to be saturated and has the lowest semi-saturation period compared with deaths and cured cases for both COVID-19 and SARS. All the three types of SARS cases are later to saturate and have longer semi-saturation period than that of COVID-19. Despite the virus caused SARS (SARS-CoV) and the virus caused COVID-19 (SARS-CoV-2) are homologous, the duration of the outbreak would be shorter for COVID-19.

## Introduction

In December 2019, a novel coronavirus-infected pneumonia causing human infection was first identified in Wuhan, a city of Hubei province located in central China with a population of 11 million people^1,2^. The coronavirus was firstly named as 2019-nCoV on January 12, 2020 and may have a probable bat origin^3^. Human to human transmission through 2019-nCoV has been confirmed among closed contacts since the middle of December 2019^4,5^. As of February 29, 2020, infections of 2019-nCoV have been identified in all Chinese provincial administrative regions and in dozens of countries around the world^6-10^. There have been a total of 79824 confirmed cases of 2019-nCoV infections in mainland China, including 2870 deaths and 41625 cases cured^11^. So far, the coronavirus has been renamed as COVID-19 on February 12, 2020 officially. Different from a previous famous beta coronavirus in 2003, severe acute respiratory syndrome coronavirus (SARS-CoV)^12-14^ that has a mortality rates of 10%^15^, that of COVID-19 is 4.13% in Hubei province, and 0.84% in rest of mainland China^11^ up to February 29, 2020.

Here, we used a descriptive model to fit data on the 2019-nCoV cases (infection, death, cure) from January 20, 2020 to February 29, 2020, aiming to capture the epidemiologic characteristics and dynamics of COVID-19 in 31 provinces in mainland China. Furthermore, we compared these characteristics with SARS in 2003 using the same model structure in order to validate this model and understand the differences among these two diseases.

## Results

### Intrinsic growth rules of COVID-19 infections in mainland China

Hubei province contributed a majority of infections to this statistical numbers (Fig 1A). Based on the cumulative infected case data of COVID-19, we firstly established a descriptive model with three parameters for each of the 31 provinces in mainland China (Fig 1B & Fig 2B, See Methods). Fitting goodness of models was expressed by the R-square, which ranged from 0.95 to 0.999, indicating that the models we established fitted the data well. The panel B of Figure 1 illustrates examples of time series data of infected cases and corresponding fitting curves in five provinces (provinces of Xinjiang, Beijing, Guangdong, Zhejiang and Heilongjiang) and that of all provinces can be found in Supplementary Figure S1. It is clear that different province varied in patterns of increase of infected cases (Fig 1B).

**Figure 1.**
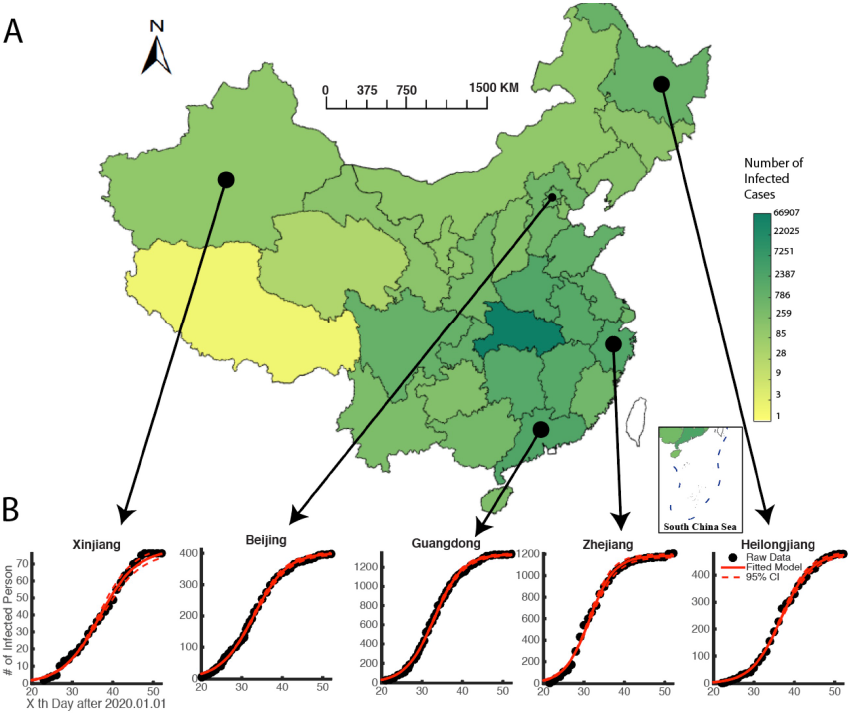
Spatial distribution of the number of COVID-19 infected cases in mainland China. Panel A is the spatial distribution of the number of COVID-19 infected cases of provinces in mainland China. Panel B takes five provinces (Xinjiang, Beijing, Guangdong, Zhejiang and Heilongjiang) as example to show the time series of infected cases and corresponding fitted curve. The horizontal axis is the *x*_th_ day after Jan 1, 2020. And the vertical axis denotes the number of infected cases in corresponding province. The black dots in each panel are raw data of the cumulative infected cases. The red line is the fitted curve by our descriptive model. The dashed red lines show the 95% confidence interval of the fitted curves.

**Fig 2.**
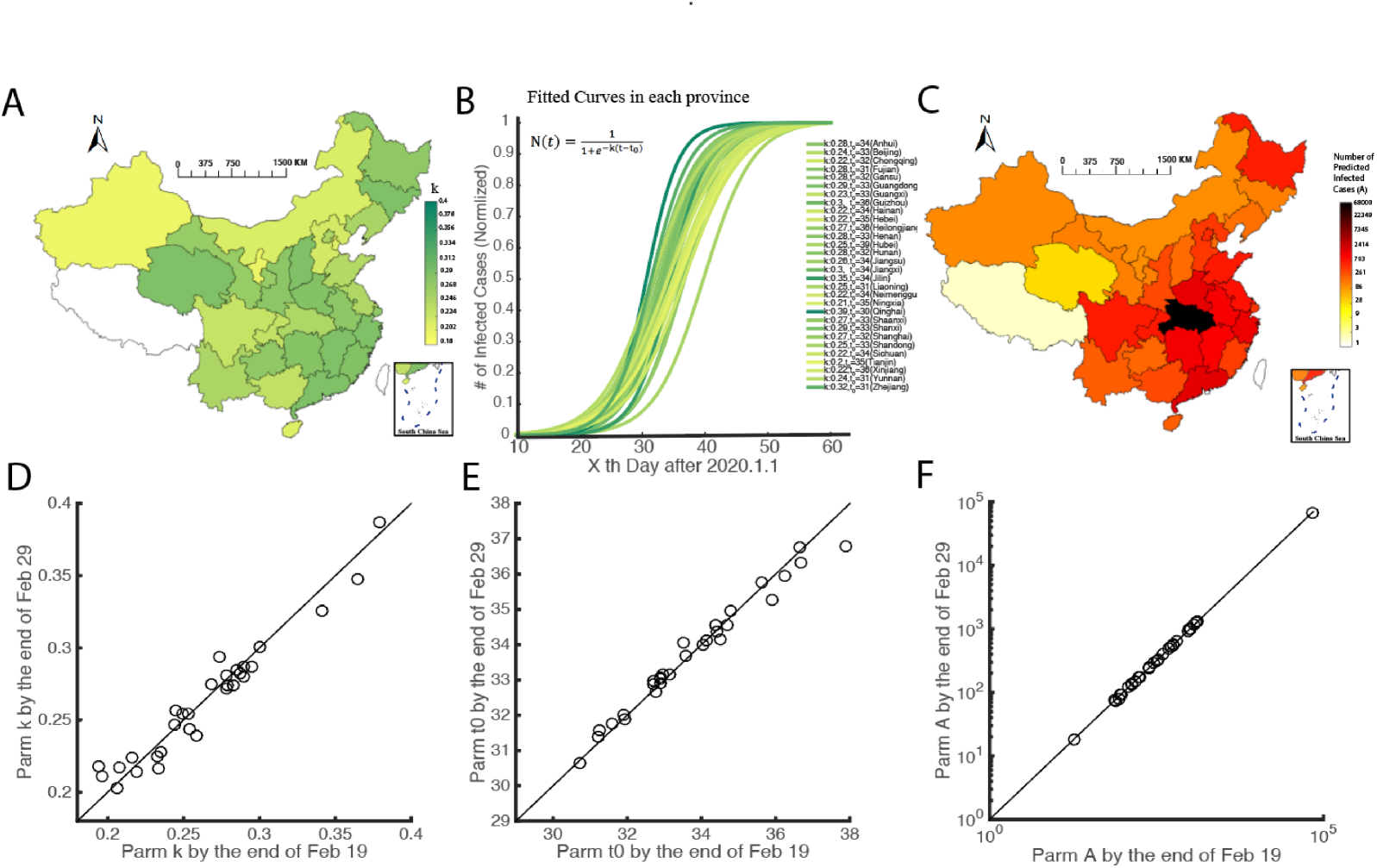
Spatial distribution of model parameters and robustness of the model. Panel A demonstrates the spatial distribution of parameter *k* of provinces in mainland China (exclude Tibet, same for B-F). Panel B shows the fitted curves of infected cases in 30 provinces (exclude Tibet). The vertical axis is the number of infected cases (normalized by dividing the maximum number of infected cases in each province). The color of each province is consistent with Panel A. Panel C is the spatial distribution of the predicted maximum number (parameter A in our model) of COVID-19 infected cases of provinces in mainland China. Panel D-F show the difference of parameters estimated from long (by the end of Feb 29) and short (by the end of Feb 29) time series data, each dot denotes for one province.

In our model, parameter *k* is an important parameter to depict the intrinsic growth rule of cumulative number of infected patients (Equation (8)). We assume that the growth pattern follows a sigmoid form, and the parameter *k* here denotes how rapid the number of infected cases get to saturate. If *k* is large, the *S* form of the curve would be sharp, and the curve will rise up very quickly to be saturated to the maximum number. While if *k* is only slightly larger than 0, the curve would show a slow and smooth increase from 0 to the maximum.

To further illustrate the spatial distribution of the infected cases’ growth pattern, we exerted further analyses of the fitted infected case curves of 30 provinces. We excluded Tibet in this analysis because the number of confirmed infected case did not increase since Jan 29, 2020. The only one confirmed case in Tibet has been cured on Feb 12. The spatial distribution of parameter *k* is shown in Fig 2A, illustrating provinces varied in the increase patterns of cumulative infected cases. Fitted curves of provinces such as Jilin and Qinghai has larger value of *k*, which means that the number of infections peaked earlier in these regions than in other provinces. By contrast, provinces like Hainan, Hebei, Ningxia, has smaller value of *k* than other provinces, suggesting the infections of these provinces would continue increasing when infections of other provinces reach saturation (Fig 2B). Fig 2C shows the spatial distribution of predicted maximum number of infected cases (parameter *A*) in each province of mainland China. With the data up to Feb. 29^th^, our model shows that the maximum number of infected COVID-19 cases in mainland China would be 80119 (95% confidence interval: [79133, 81105]). Among all the provinces in mainland China, the Hubei province is predicted to have largest number of infected COVID-19 cases, which would be 66750 (95% confidence interval: [65939,67561]).

Our model could not only well explain the data in various geographic space, but also provided robust results with different length of time series (Fig 2D-F). We used our model to fit the data by Feb 19 and Feb 29 respectively, and the parameters estimated by the two dataset do not have significant difference (for k, t=0.09, p=0.93; for t_0_, t=0.04, p=0.97; for A, t=0.02, p=0.98; two-tailed test). This model’s spatial and temporal robustness provides us confidence in further analyses.

## Comparison of intrinsic epidemic rules between COVID-19 in Wuhan and SARS in Beijing (2003)

To compare the intrinsic growth rule of infected cases, dead cases and cured cases between COVID-19 and SARS, we applied the descriptive model to fit the data of SARS in Beijing 2003 (Fig 3A). Since the time series data of SARS was full phase of the outbreak, the parameters estimated by our model was solid (R-square>0.95). We found the *k* value of fitted curves of infected SARS cases in Beijing was 0.18, which was much lower than that of COVID-19 (*k* = 0.25) in Hubei Province (Fig 3 ABC). Furthermore, we found that all 30 provinces’ *k* values were significantly larger than 0.18 (t=10.71, p<0.0001, right-tailed test) (Fig 3D). We also noted the *t*_*0*_ of fitted curves of infected SARS cases in Beijing was larger than that of fitted curves of infected COVID-19 cases in all provinces (Fig 3E). Moreover, the increasing rate (*k*) was negatively correlated (Pearson, *r* = −0.44, *p* = 0.0125) to semi-saturation period (Fig 3F). We concluded that at the provincial level, the COVID-19 spread more quickly than the SARS did. The infections of COVID-19 increased more rapid and would reach saturation earlier than SARS according to our model.

**Fig 3.**
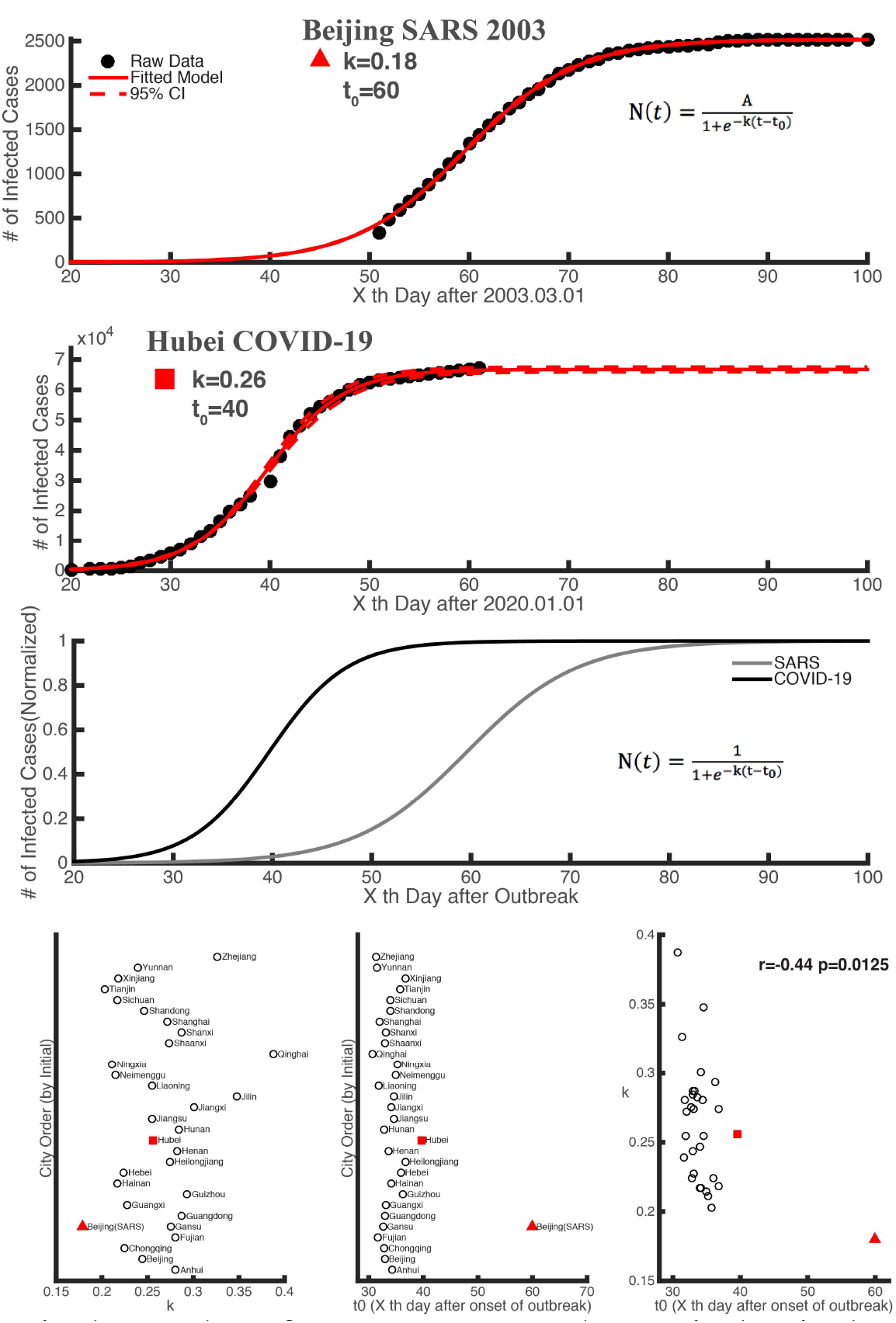
Comparison of intrinsic epidemic rules between COVID-19 in Wuhan and SARS in Beijing (2003) Panel A shows the raw data and fitted curve of SARS cases in Beijing. The horizontal axis is the x^th^ day after March 1, 2003. The vertical axis denotes the cumulative number of SARS cases in Beijing. The black dots are the raw data of the SARS cases. The red line is the fitted curve by our descriptive model. The dashed red lines are the 95% confidence interval of the fitted curves. Panel B shows the raw data and fitted curve of COVID-19 cases in Hubei province. The horizontal axis is the x_th_ day after Jan 1, 2020. The vertical axis denotes the cumulative number of COVID-19 cases in Hubei. The black dots are raw data of the COVID-19 cases. The red line is the fitted curve by our descriptive model. The dashed red lines are the 95% confidence interval of the fitted curves. Pannel C shows the fitted curves of infected SARS cases (gray curve) and COVID-19 (black curve), which are normalized by dividing the maximum number of infected cases in each province. Panel D illustrates the *k* value (increasing rate of infected cases) of fitted curves of infected COVID-19 cases in each province in mainland China, where Hubei is represented by the solid red square (the same in D and E). *k* value of fitted curves of infected SARS cases in Beijing is represented by the solid red triangle (the same in D and E). Panel E illustrates the *t*_*0*_ (semi-saturation period) of fitted curves of infected COVID-19 cases in each province in mainland China, as well as *t*_*0*_ of fitted curves of infected SARS cases in Beijing. The scatter plot of *k* and *t*_*0*_ is shown in Panel F, indicating significant negative correlation between *k* and *t*_*0*._

## Intrinsic rules of infection, death and recovery of COVID-19 and SARS2003

The outbreak of SARS in Beijing was the most serious among all the provincial administrative regions, and the outbreak of COVID-19 was most serious in Hubei province. Here we used our descriptive model to fit the dead and cured cases’ curves for SARS in Beijing and COVID-19 in Hubei province (Fig 4 A-C, F-H, R-square ≥ 0.95, for other provinces in mainland China, see Fig S2, S3), and compared the growth features of three types cases of SARS and COVID-19 by two parameters, *k* and *t*_*0*_ (Fig 4 DEIJ). We found that *k* values of the three types cases of SARS were lower than that of COVID-19 (Fig 4 DE), and semi-saturation period for them in SARS was higher than COVID-19’s, which suggested the duration of COVID-19 would be shorter than that of SARS. For both SARS and COVID-19, their infected curves have the lowest semi-saturation period (Fig 4 EJ), while cured curves have the highest which is in keeping with the common sense that the cure is always later than infection and death. For both SARS in Beijing and COVID-19 in Hubei, the *k* values of death case curves were lowest compared with curves of infected cases and cured cases. For both SARS and COVID-19, the *k* value of infected case curve was larger than that of cured curve.

**Fig 4.**
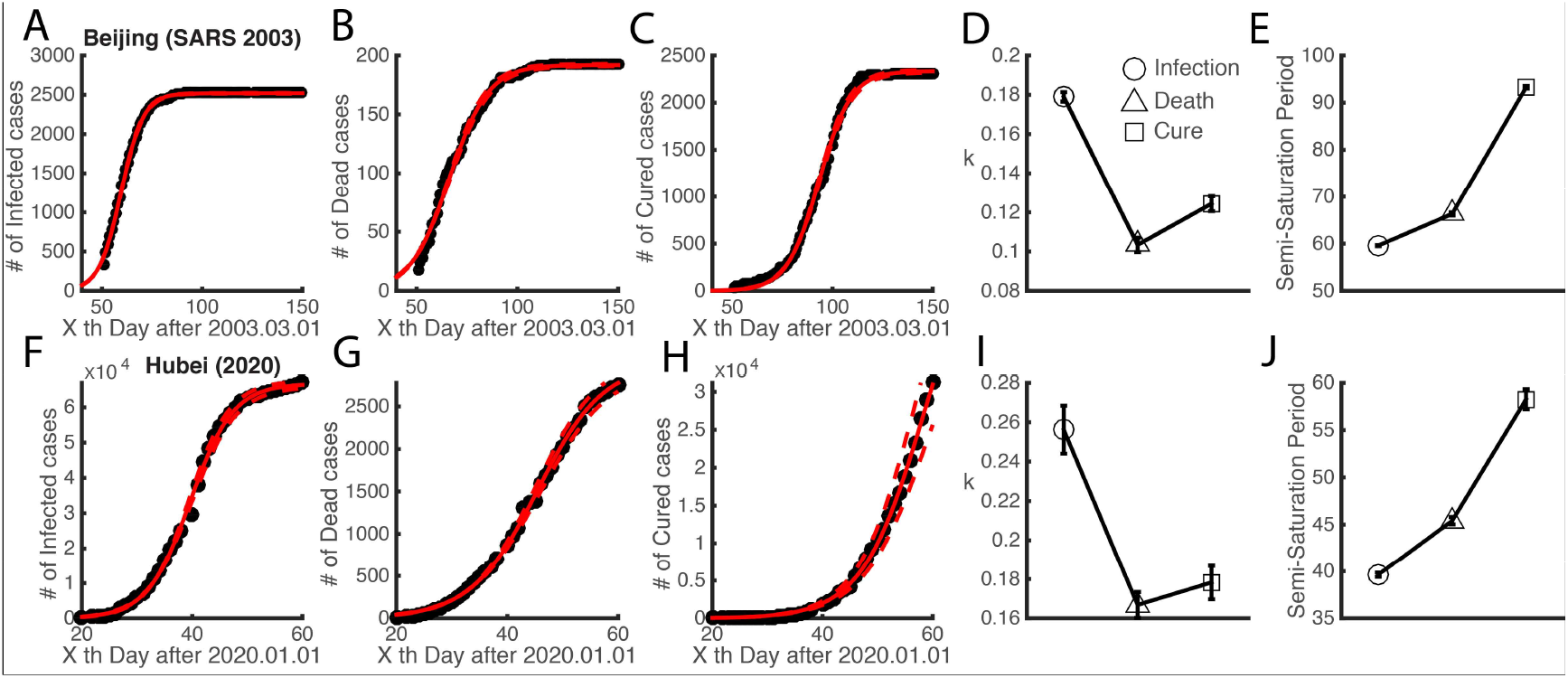
Intrinsic rules of infection, death and recovery of COVID-19 and SARS2003. Panel A-C show the raw data and fitted curve of SARS cases in Beijing. Their horizontal axises are the x_th_ day after Mar 1, 2003. The black dots in Panel A, B and C are raw data of the SARS infected cases, dead cases and cured cases, respectively. The red lines are the fitted curves by our descriptive model. The dashed red lines are the 95% confidence interval of the fitted curves. Panel D and Panel E respectively shows the *k* value and the *t*_*0*_ of fitted curves of three types cases caused by SARS (the circle, triangle and square denotes infected, dead and cured cases, respectively, and the same in Panel I and J). The error bars show the 95% confidence interval of the parameters. Panel F-H show the raw data and fitted curve of COVID-19 cases in Hubei. Their horizontal axises are the x_th_ day after Jan 1, 2020. The black dots in Panel F, G and H are raw data of the COVID-19 infected cases, dead cases and cured cases, respectively. The red lines are the fitted curves by our descriptive model. The dashed red lines are the 95% confidence interval of the fitted curves. Panel I and Panel J respectively shows the *k* value and the *t*_*0*_ of fitted curves of three types cases caused by COVID-19, which is same with D and E.

## Discussion

We modeled the epidemiologic characteristics of 2019-nCoV for 31 provinces in mainland China based on the data from Jan 10 to Feb 29 during the outbreak (Hubei province reported laboratory confirmed 2019-nCoV cases since Jan 10, while other provinces in mainland China reported laboratory confirmed 2019-nCoV cases since Jan 20). Furthermore, we compared these characteristics with SARS in 2003 using the same model framework.

Compared with dynamical models such as SIR (Susceptible - Infectious - Recovered) or SEIR (Susceptible - Exposed - Infectious - Recovered), our model is relatively simple but robust. We used a differential equation in logistic form to describe the dynamics of the infected, dead and healed cases caused by COVID-19 respectively (See Methods). The increasing of the population no matter in which type of case (infection, death or recovery) has multiple reasons. When the outbreak of COVID-19 originated in Wuhan, it did not cause too much concern at the beginning so that it leads to many infections and starts to export potential exposed cases to other provinces. But as the successive closure of the cities in Hubei province, and isolation of the community in all the provinces in mainland China, the virus spread tendency was weakened. In our model, we also considered these measures taken by the government. We rewrote Equation (1) to Equation (9).

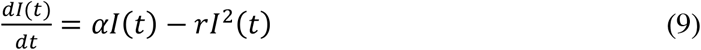

Where *α* is growth rate of the number of infections; *I(t)* is cumulative number of infections at time *t*; *r* is a parameter to reflect the effect of measures aiming at controlling the virus spread. Here we decompose *I (t)* into two parts. The first part *αI*(t) represents the rapid infections growth in the early phase of the outbreak without any measures for epidemic control. The second part *rI*^2^(t) illustrates the slower infections growth after taking effective isolation measures for epidemic control. For death cases, this idea still holds. But things are different for cured case. In contrast, at the early phase of the outbreak, the number of cured cases increase slowly. But once there are effective treatments, the number of cured cases will rise up quickly. The growth of cured cases would again be slow since the number of patients is limited. All these facts were considered in our highly summarized model (Equation (8)) in a macroscopic scale. Each fitted model has a *R*-square value greater than 0.95 to the original data set, so that we could further discuss some intrinsic mechanisms for the growth rules of infection, death and recovery through the parameters estimated from this model (See Methods).

The value of *k* from Equation (8) is a critical parameter. *k* represents how rapid the number of infected cases get to the saturation. We found the variability of *k* values among fitted curves of infected COVID-19 cases in 30 provinces (exclude Tibet) (Fig 2 AB) is large (Ranging from 0.19 to 0.38). It is clear to show the real situation in each province and this guides us to be more careful against the epidemic. Further we compared this characteristics between COVID-19 and SARS in 2003 in typical provinces (Fig 3 AB). We noted that *k* values of fitted COVID-19 infection curves in 30 provinces of mainland China (exclude Tibet) are larger than that of SARS in Beijing 2003 (Fig 3 C), which suggests the time cost of this epidemic will be shorter compared with SARS in 2003. This result may relate to several aspects. Firstly, it may due to the basic spread property of the virus. Besides, the isolation policy of all the provinces in mainland China was more strictly enforced than it was in 2003. Moreover, the improved clinical skills today may also helpful for shortening the duration of epidemic.

Furthermore, we analyzed semi-saturation period (Fig 4 EK) (*t*_*0*_ in Equation (8)) and make comparison in three types of cases (infection, death and recovery) (Fig 4 A-C G-I). *k* values for the fitted curves of the three types of cases caused by COVID-19 were larger than that of SARS, while *t*_*0*_ values were the opposite. This again suggests that the epidemic of COVID-19 will be quicker to pass away than SARS. For both SARS and COVID-19, fitted curves of infections have lowest semi-saturation period (Fig 4 EK), while fitted curves of cured cases have the highest semi-saturation period, which follows the common sense that the cure was always later than infection and death since it costs time to get on the research of effect plans to cure the patients. For both SARS and COVID-19, the *k* value of infected case curve was larger than that of cured curve, while that of death case is the lowest.

## Methods Sources of data

We obtained the time series data of 2019-nCoV cases from the National Health Commission of China, and the provincial Health Commission of 31 provincial administrative regions in mainland China (January 10 to February 29, 2020). The data used in this study include the number of confirmed 2019-nCoV cases, the cumulative number of suspected, dead, and cured cases. The 200 infected cases found in jail of Shandong Province on Feb 20 were not counted in. All cases were laboratory confirmed following the case definition by national health commission of China^16^. The basic test procedure has been described in detail in previous work^4,17^.

The severe acute respiratory syndrome (SARS) in 2003 was another fatal coronavirus epidemics over the last two decades, we collected the data (the cumulative number of suspected, dead, and cured cases) of SARS during March 1 to August 16, 2003, in Beijing from the National Health and Family Planning Commission of PRC.

## Epidemic curve modeling

We assumed the number of susceptible people per unit time was proportional to the number of uninfected persons, and isolation or closure of the city could affect the spread effect of infected to susceptible people to a certain extent. Previous studies have showed that daily cumulative number of patients could be explained by logistic function^18,19^. Based on the above assumption and existing findings, we modeled the epidemic information of virus as logistic form (Equation (1))

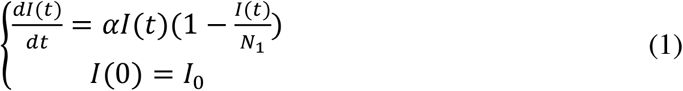

where *N*_*1*_ is maximum number of cumulative infections, *I(t)* is cumulative number of patients at time *t, α* is the incidence growth rate, and *I*_*0*_ is the number of infected cases at the initial time.

Analytical solution of Equation (1) could be written in the form of Equation (2)

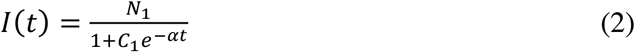

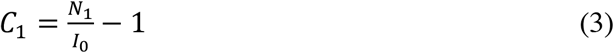

Similarly, we could write the analytical solutions for the dead and cured cases by

Equation (4) and Equation (6).

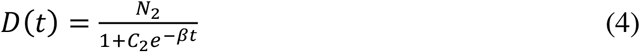

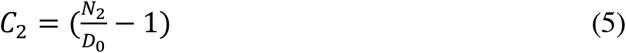

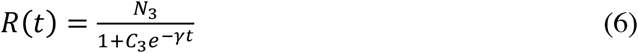

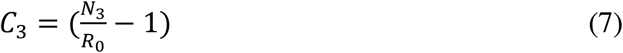

Where *β, γ* is the growth rate of dead cases and the growth rate of cured cases, respectively. *N*_*2*_ and *N*_*3*_ is maximum number of cumulative dead and cured cases. *D*_*0*_ and *R*_*0*_ is the number of dead and cured cases at initial time.

Here we simplified Equation (2),(4),(6) and unified them as Equation (8)

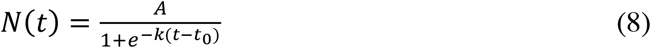

where *N(t)* is the general form of *I(t), D(t)* or *R(t), A* denotes *N*_*1*_, *N*_*2*_, *or N*_*3*_, and *k* represents *α, β*, or *γ*, indicating the intrinsic rule for growth of cases. *t*_*0*_ represents the semi-saturation period, illustrating the inflection point of the sigmoid curve.

The data processing and modeling were performed on MATLAB (The MathWorks) with custom scripts. The nonlinear least square (NLS) algorithm was adopted for data fitting and parameter estimation. We used the MATLAB function “nlinfit” to minimize the sum of squared differences between the data points and the fitted values. Equation (8) was used in this work as the descriptive model to depict the intrinsic growth rule for infected, dead and cured cases.

## Data Availability

Data is obtained from the open website of the health commissions in 31 provinces in mainland China

## Acknowledgements

This study is sponsored by the National Key Research and Development Program of China (2018YFC1508903), the National Natural Science Foundation of China (41621061) and the support of International Center for Collaborative Research on Disaster Risk Reduction (ICCR-DRR).

## ADDITIONAL INFORMATION

### Contributions

C.L.H, Y.M.L and S.N.Y designed the research. C.L.H and Y.M.L performed the research. C.L.H and Y.M.L. analyzed the data. Y.M.L, C.L.H and S.N.Y wrote the paper.

### Competing interests

The authors declare no competing interests.

**Supplement Figure S1.**
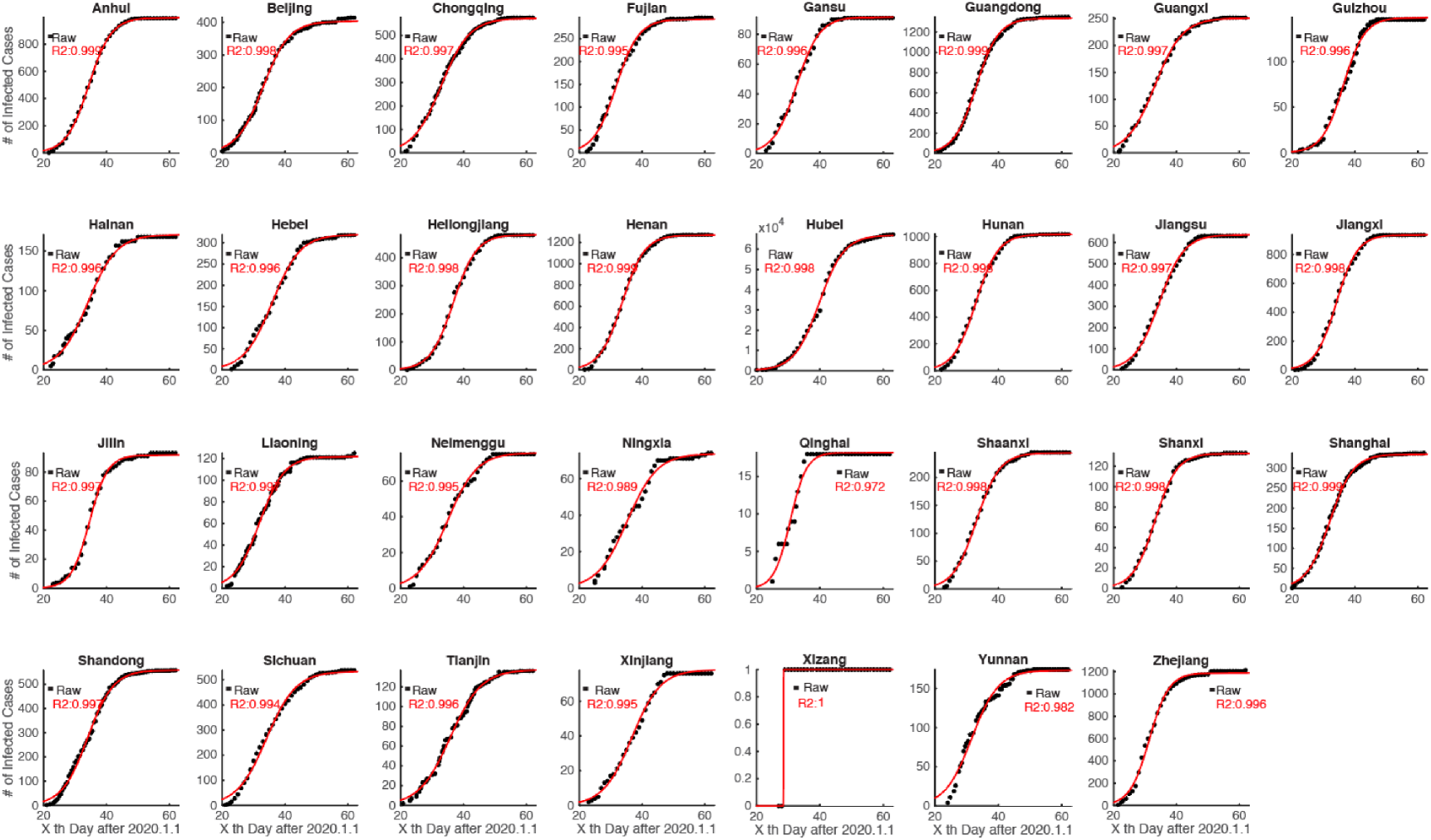
Intrinsic growth rules of patients infected with 2019 novel coronavirus in all provinces of mainland China. The black dots in each panel are raw data of the cumulative infected cases. The red line is the fitted curve by our descriptive model.

**Supplement Figure S2.**
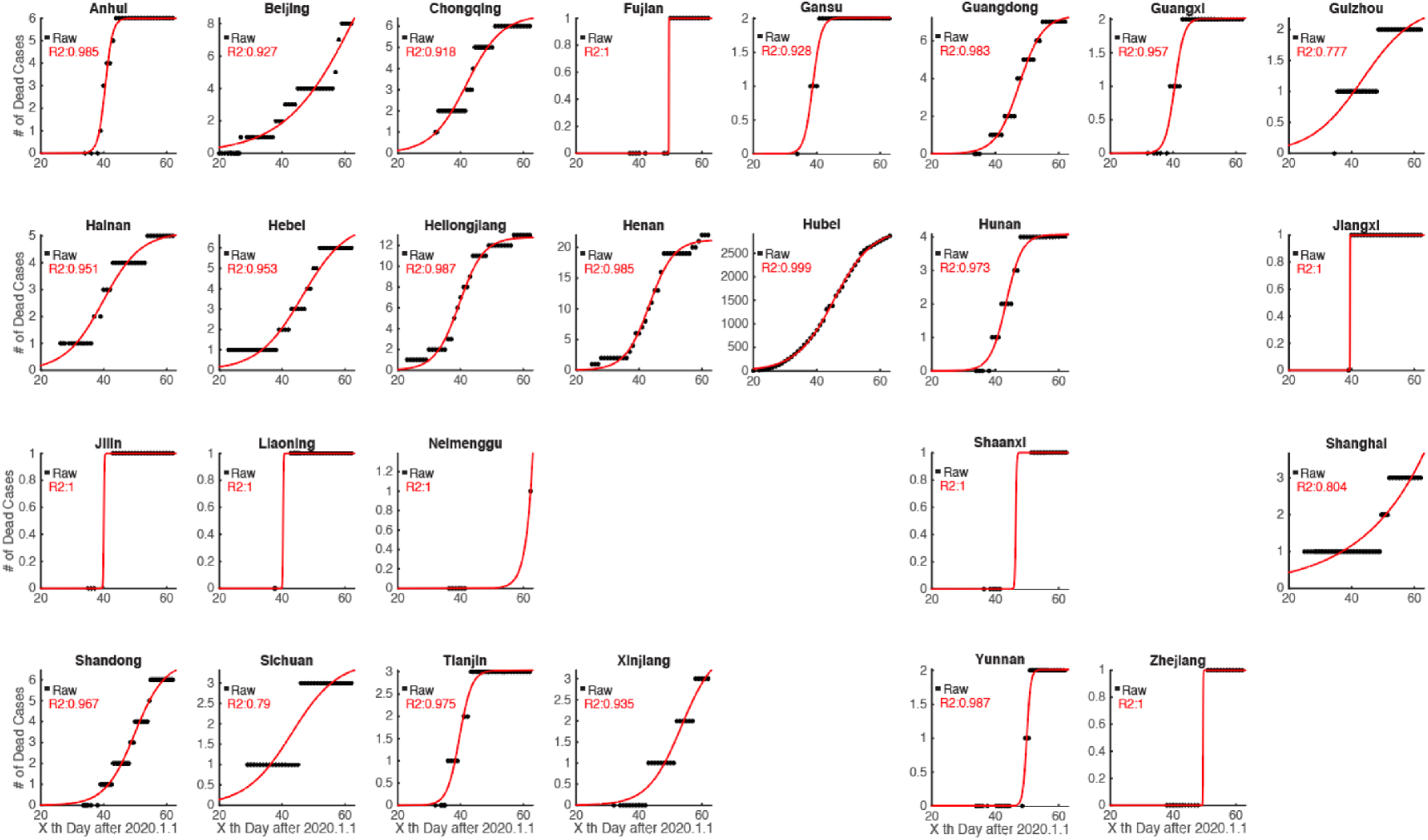
Intrinsic growth rules of patients dead with 2019 novel coronavirus in all provinces of mainland China (Excluded 0 death case provinces) The black dots in each panel are raw data of the cumulative dead cases. The red line is the fitted curve by our descriptive model.

**Supplement Figure S3.**
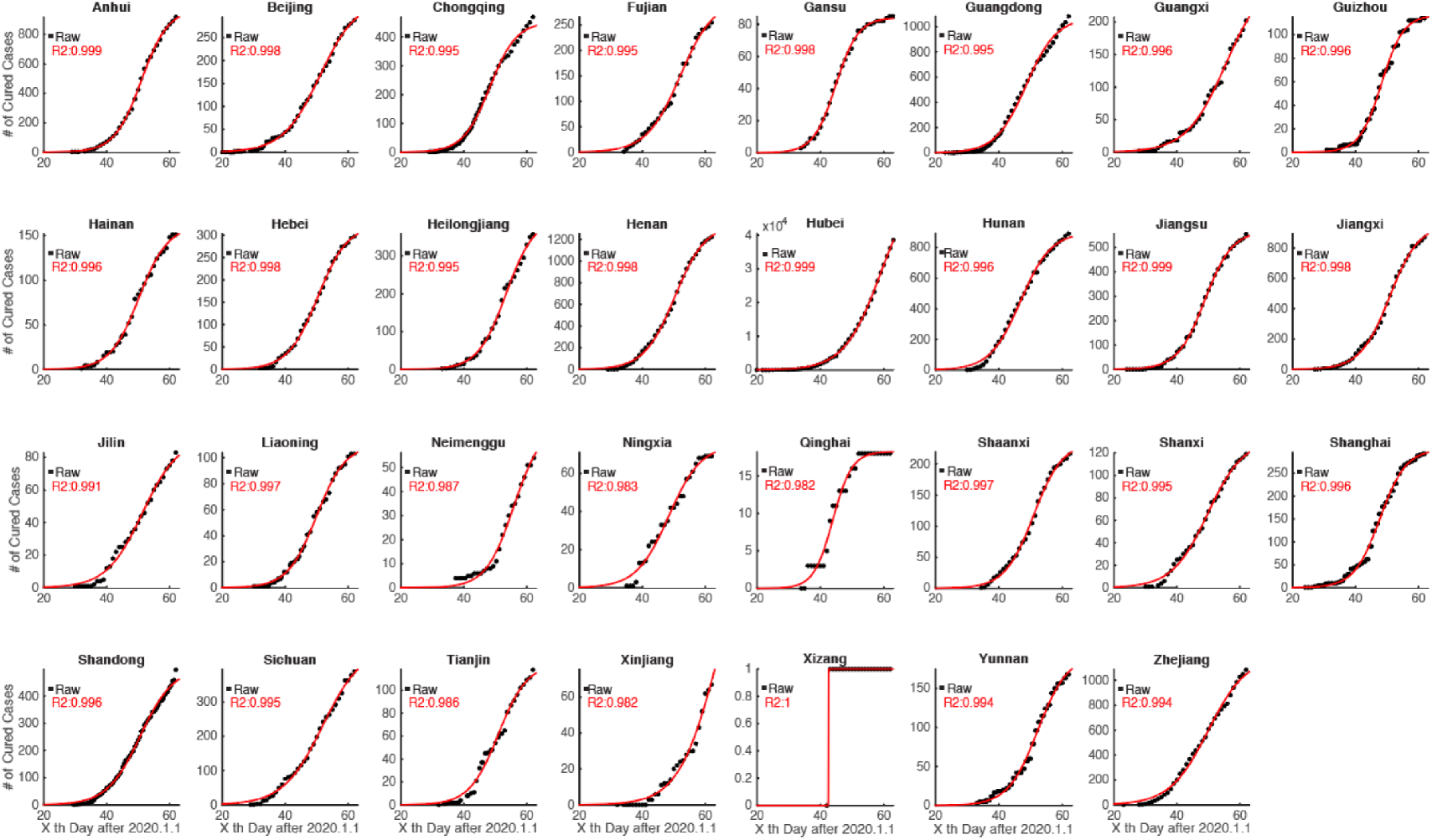
Intrinsic growth rules of patients cured with 2019 novel coronavirus in all provinces of mainland China. The black dots in each panel are raw data of the cumulative cured cases. The red line is the fitted curve by our descriptive model.

## References

1 Zhu, N. et al.. A Novel Coronavirus from Patients with Pneumonia in China, 2019. N Engl J Med 382, 727–733, doi:10.1056/NEJMoa2001017 (2020).

2 Hui, D. S. et al.. The continuing 2019-nCoV epidemic threat of novel coronaviruses to global health - The latest 2019 novel coronavirus outbreak in Wuhan, China. Int J Infect Dis 91, 264–266, doi:10.1016/j.ijid.2020.01.009 (2020).

3 Zhou, P. et al.. A pneumonia outbreak associated with a new coronavirus of probable bat origin. Nature, doi:10.1038/s41586-020-2012-7 (2020).

4 Li, Q. et al.. Early Transmission Dynamics in Wuhan, China, of Novel Coronavirus-Infected Pneumonia. N Engl J Med, doi:10.1056/NEJMoa2001316 (2020).

5 Wang, D. et al.. Clinical Characteristics of 138 Hospitalized Patients With 2019 Novel Coronavirus-Infected Pneumonia in Wuhan, China. JAMA, doi:10.1001/jama.2020.1585 (2020).

6 Holshue, M. L. et al.. First Case of 2019 Novel Coronavirus in the United States. N Engl J Med, doi:10.1056/NEJMoa2001191 (2020).

7 Ki, M. & nCo, V. T. Epidemiologic characteristics of early cases with 2019 novel coronavirus (2019-nCoV) disease in Republic of Korea. Epidemiol Health, e2020007, doi:10.4178/epih.e2020007 (2020).

8 Shigemura, J., Ursano, R. J., Morganstein, J. C., Kurosawa, M. & Benedek, D.M. Public responses to the novel 2019 coronavirus (2019-nCoV) in Japan: Mental health consequences and target populations. Psychiatry Clin Neurosci, doi:10.1111/pcn.12988 (2020).

9 Munster, V. J., Koopmans, M., van Doremalen, N., van Riel, D. & de Wit, E. A Novel Coronavirus Emerging in China - Key Questions for Impact Assessment. N Engl J Med 382, 692–694, doi:10.1056/NEJMp2000929 (2020).

10 Nishiura, H. et al.. The Rate of Underascertainment of Novel Coronavirus (2019-nCoV) Infection: Estimation Using Japanese Passengers Data on Evacuation Flights. J Clin Med 9, doi:10.3390/jcm9020419 (2020).

11 Prevention, C. C. f. D. C. a. The latest situation of COVID-19,< http://www.nhc.gov.cn/xcs/yqtb/202003/9d462194284840ad96ce75eb8e4c8039.shtml> (2020).

12 Drosten, C. et al.. Identification of a novel coronavirus in patients with severe acute respiratory syndrome. New Engl J Med 348, 1967–1976, doi: DOI 10.1056/NEJMoa030747 (2003).

13 Ksiazek, T. G. et al.. A novel coronavirus associated with severe acute respiratory syndrome. N Engl J Med 348, 1953–1966, doi:10.1056/NEJMoa030781 (2003).

14 Kuiken, T. et al.. Newly discovered coronavirus as the primary cause of severe acute respiratory syndrome. Lancet 362, 263–270, doi: Doi. 10.1016/S0140-6736(03)13967-0 (2003).

15 WHO. Summary of probable SARS cases with onset of illness from 1 November 2002 to 31 July 2003,<https://www.who.int/csr/sars/country/table2004_04_21/en/> (2003).

16 Prevention, C. C. f. D. C. a. Prevention and control measures of COVID-19,<http://www.nhc.gov.cn/jkj/s3577/202002/a5d6f7b8c48c451c87dba14889b30147/files/3514cb996ae24e2faf65953b4ecd0df4.pdf > (2020).

17 Huang, C. et al.. Clinical features of patients infected with 2019 novel coronavirus in Wuhan, China. Lancet 395, 497–506, doi:10.1016/S0140-6736(20)30183-5 (2020).

18 Huang, D., Guan, P. & Zhou, B. Fitness of morbidity and discussion of epidemic characteristics of SARS based on logistic models. Chinese Journal of Public Health 19, 1–2 (2003).

19 Wang, Y. & Liu, x. The compound Logistic model used to describe epidemic situation dynamics of SARS in Beijing. Journal of China Jiliang University 16, 159–162 (2005).

